# Pooled Sample Testing for SARS-CoV-2 Using Rapid RT-PCR COVID-19 Tests

**DOI:** 10.1101/2021.01.22.21250339

**Authors:** Bethany Hyde, Prat Verma, Ethan M. Berke

**Affiliations:** UnitedHealth Group, Minnetonka, MN, USA

**Keywords:** COVID-19, testing, PCR, polymerase chain reaction, pooled samples, SARS-CoV-2

## Abstract

We tested an operationally efficient way to pool samples on a rapid, point-of-care PCR device and examined the limit of detection of SARS-CoV-2 for various pool sizes. Pooled testing maintained testing performance similar to individual sample PCR testing, offering the potential for scalable rapid testing at lower cost with less supplies.

---

Given the often asymptomatic presentation of COVID-19 and the potential for viral spread before the onset of symptoms, reliable and repeatable testing is necessary to slow the pandemic. New models of testing are required that can be self-administered, offer rapid results, and be deployed in a variety of settings. Pooled sampling allows for cost-effective testing at improved scale. Samples are combined and jointly tested with a single lab test. If the test is negative, all samples are considered negative. If the test is positive, each sample may be tested individually to determine which sample is positive.

Abdalhamid et al. found that for communities with a 5% prevalence rate, pooling five samples in one test provided a 57% reduction in the total number of tests needed compared to individual testing.(*1*) Lohse et al. found that up to 30 samples could be combined while preserving diagnostic accuracy.(*2*)

Using pooled samples on a point-of-care device allows for the additional advantage of rapid result turnaround and simplification of logistics, allowing for isolation decisions to be made in real time. To advance this concept, we tested an operationally efficient way to pool samples on a rapid-resulting PCR device and examined the limit of detection of SARS-CoV-2 for various pool sizes on a point-of-care test.

## The Study

Optum Labs, the research and development unit at UnitedHealth Group, performed this research under an IRB approved protocol. We held testing events over the course of several weeks where asymptomatic employees were tested. All participants validated they were not experiencing any symptoms at the time of testing using both a daily symptom-checking application, ProtectWell, plus a symptom check at time of testing.

We determined pool sizes based on the number of participants and the number of swabs each participant provided, using a convenience sample approach. Participants provided 1–3 self-collected anterior nares swabs, from both nostrils. For a given pool, each individual participant swab was inserted in a single dilution buffer tube, provided with the test, for the pool and agitated for a minimum of 10 seconds. Subsequent individual swabs were similarly exposed to dilution buffer until the pool contained all samples. We used a portable, rapid RT-PCR device (Visby Medical, San Jose, CA) to test all pools per manufacturer instructions.

A second swab collected at the same time was run individually for each participant on either another Visby RT-PCR device or BioRad RT-PCR to ascertain the status of each member of the pool. All participants were negative for SARS-CoV-2.

Using the remaining dilution solution from the pools, we calculated a target viral load of the sample based on remaining volume and added various concentrations of positive control (ZeptoMetrix NATSARS(COV2)-ERC1) to the dilution solution. Positive control was added to the sample with a lab pipette to ensure accurate measurement, and a repeat pooled test was performed with the original negative participants plus the viral control.

The portable RT-PCR device detected low levels of positive control in pools of up to 15 participants (Table 1). For pools larger than 15 participants, the device was less consistent unless the viral copies/mL were increased. However, at these large pool sizes the device detected presence of virus at 9,483 viral copies per milliliter—lower than the limit of detection for many COVID-19 tests approved by the FDA, and in the performance range of some approved RT-PCR methods.

We created one additional pool with a known positive participant’s sample combined with 14 other participants who were known negative. This pool was positive. The positive participant was 12 days from exposure and substantially recovered from COVID-19. Interestingly, the individual’s sample was read as negative on a BioRad PCR machine, which has a published limit of detection of 10,000 viral copies/mL.

## Conclusions

This study validates that pooled testing is a compelling way to increase testing capacity while still using high performance testing technology. Further analysis is needed to clearly define the limit of detection in pools greater than 15 participants.

This study had several limitations. Pool sizes were opportunistic rather than formulaic. In most pools, positive control was added via a pipette to ensure exact measurements, but in a few limited cases the amount of positive control was approximated.

While the dilution buffer can preserve a sample for approximately two hours before degrading, in a few cases the sample may have been processed up to an hour after collection (most samples were processed in 30 minutes or less). Our observations support that testing as soon as possible after introduction into the dilution buffer increases the performance of the test, particularly in detecting low viral loads in larger pools.

Despite these limitations, this work demonstrates the potential of pooled testing with pool sizes of 15 more to improve operational efficiency while reducing cost and supplies and maintaining testing performance similar to other PCR testing approaches.

## Data Availability

Data is summarized in the table included in the manuscript.

## Author Bio

(first author only, unless there are only 2 authors)

Bethany Hyde, MHA is a director of research and development at Optum Labs, where she supports a variety of research initiatives including COVID-19 testing, sexually transmitted infections, and chronic kidney disease.

---

Comparison of COVID-19 pooled samples and viral copies per mL.

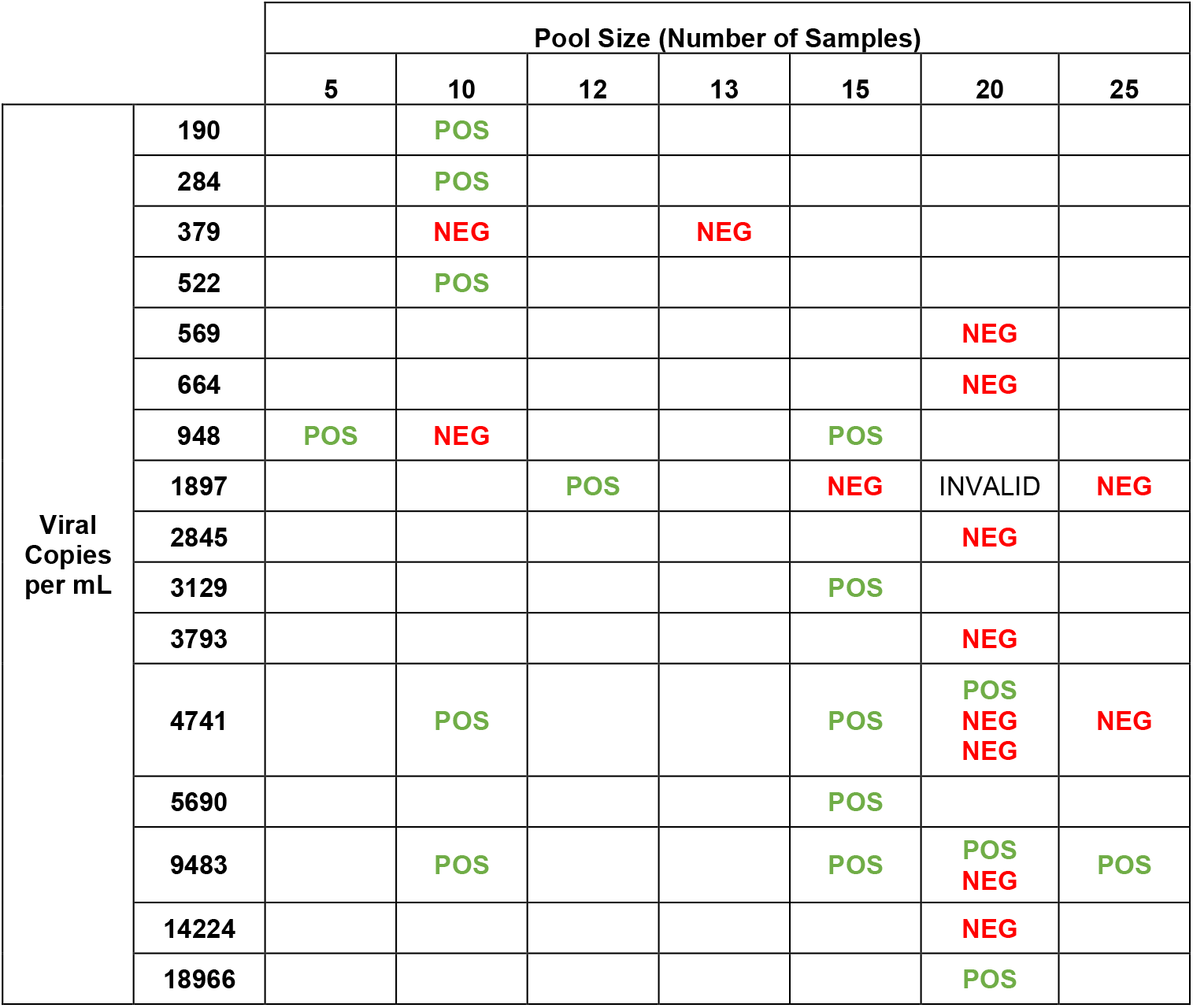

|  |  | Pool Size (Number of Samples) |  |  |  |  |  |  |
| --- | --- | --- | --- | --- | --- | --- | --- | --- |
|  |  | 5 | 10 | 12 | 13 | 15 | 20 | 25 |
| Viral<br>Copies<br>per mL | 190 |  | POS |  |  |  |  |  |
|  | 284 |  | POS |  |  |  |  |  |
|  | 379 |  | NEG |  | NEG |  |  |  |
|  | 522 |  | POS |  |  |  |  |  |
|  | 569 |  |  |  |  |  | NEG |  |
|  | 664 |  |  |  |  |  | NEG |  |
|  | 948 | POS | NEG |  |  | POS |  |  |
|  | 1897 |  |  | POS |  | NEG | INVALID | NEG |
|  | 2845 |  |  |  |  |  | NEG |  |
|  | 3129 |  |  |  |  | POS |  |  |
|  | 3793 |  |  |  |  |  | NEG |  |
|  | 4741 |  | POS |  |  | POS | POS<br>NEG<br>NEG | NEG |
|  | 5690 |  |  |  |  | POS |  |  |
|  | 9483 |  | POS |  |  | POS | POS<br>NEG | POS |
|  | 14224 |  |  |  |  |  | NEG |  |
|  | 18966 |  |  |  |  |  | POS |  |

